# The effect of osteotomy technique (flat-cut vs wedge-cut Weil) on pain relief and complication incidence following surgical treatment for patients presenting with metatarsalgia in a private metropolitan clinic: Protocol for a randomised controlled trial

**DOI:** 10.1101/2020.12.10.20242339

**Authors:** Manaal Fatima, Nalan Ektas, Corey Scholes, Michael Symes, Andrew Wines

**Affiliations:** EBM Analytics; Sydney Orthopaedic Trauma and Reconstructive Surgery; North Sydney Orthopaedic and Sports Medicine Clinic

**Keywords:** Metatarsalgia, Weil osteotomy, pain, complication, surgery

## Abstract

**Introduction:** Weil osteotomies are performed to surgically treat metatarsalgia, by shortening the metatarsal via either a single distal oblique cut (flat-cut) with translation of the metatarsal head, or through removal of a slice of bone (wedge-cut). The wedge-cut technique purportedly has functional and mechanical advantages over the flat-cut procedure, however in-vivo data and quality of evidence is currently lacking. This study aims to investigate whether wedge-cut Weil osteotomy compared to traditional flat-cut Weil is associated with increased pain relief and fewer complications up to 12 months postoperatively.

**Methods and analysis:** Patient, surgical and clinical data will be collected for 80 consecutive consenting patients electing to undergo surgical treatment of propulsive metatarsalgia in a randomised control trial, embedded within a clinical registry. The primary outcome is patient-reported pain as assessed by the Foot and Ankle Outcome Score (FAOS) - Pain subscale, and the secondary outcome is the incidence of procedure-specific complications at up to 12 months postoperatively. The groups will be randomised using a central computer-based simple randomisation system, with a 1:1 allocation without blocking and allocation concealment. A mixed-effects analysis of covariance will be used to assess the primary outcome, with confounders factored into the model. A binary logistic regression will be used to assess the secondary outcome in a multivariable model containing the same confounders.

**Trial registration:** Australian New Zealand Clinical Trials Registry, ACTRN12620001251910. Registered on 23 November 2020.

**Ethics and dissemination:** Ethics approval for this study was provided through the NSW/VIC branch of the Ramsay Health Care Human Research Ethics Committee (HREC approval number 2020-007). The results of this study will be disseminated through peer-reviewed journals and conference presentations.

**Strengths and limitations of this study:**

- To the best of the authors knowledge, the trial will be the first to examine clinical efficacy of the wedge-cut Weil osteotomy compared to the flat-cut technique with a prospective, randomised control design.
- A sample size of N=80 participants will ensure adequate power to detect differences between the control and experimental groups, with an allowance for a dropout rate of 10%.
- A limitation in the methods is the routine performance of adjunct surgical procedures for lesser toe or soft tissue correction. The statistical plan aims to control for this by treating adjunct procedures as potential confounders of the effect of the intervention, along with prognostic factors identified from available literature on pain ratings in metatarsalgia. These confounders will be included in the models used to analyse the primary and secondary outcomes.

## INTRODUCTION

### Background and rationale

Propulsive metatarsalgia is defined as pain under one or more metatarsal heads during the “third rocker” phase (30% to 60%) of the gait cycle, from heel lift-off to the end of propulsion by the great toe (Besse, 2017). Weil osteotomy is a reliable distal oblique osteotomy procedure performed to treat lesser metatarsal deformities and alleviate metatarsalgia, by shortening the metatarsal in the transverse plane. The osteotomy is performed with or without adjunct procedures for toe correction, which may include proximal interphalangeal arthrodesis (Jay et al., 2016), fusion/arthrodesis of the first metatarsophalangeal (MTP) joint (Donegan & Blume, 2017), or hallux valgus correction (Bia et al., 2018). While Weil osteotomies are commonly performed, complications include floating toe, joint stiffness, avascular necrosis, transfer of metatarsalgia to subsequent toes, and plantar flexion of the metatarsal (Besse, 2017; Highlander et al., 2011; Lunz et al., 2010).

Traditional flat-cut Weil osteotomies involve a single distal oblique incision in the dorsal aspect of the metatarsal head, with translation of the head by 5-10mm. Wedge-cut osteotomy is a modification of the flat-cut Weil procedure, and includes a second incision to remove a slice of bone (Besse, 2017). A wedge is created either with parallel sides or with an apex on the plantar aspect of the metatarsal, and the procedure is purported to reduce plantar translation of the metatarsal head, maintain the MTP centre of rotation and improve intrinsic muscle function as demonstrated on sawbone models (Melamed et al., 2002). However, there is limited in-vivo data for the clinical efficacy of this technique, and the quality of evidence is lacking (Garg et al., 2008; Melamed et al., 2002).

The proposed study has therefore been designed to investigate whether the modified wedge-cut Weil osteotomy compared to the flat-cut technique, with or without required adjunct procedures, is associated with increased pain relief and fewer complications at up to 12 months postoperatively in patients presenting with propulsive metatarsalgia.

### Objectives

The primary objective of this study is to determine, in patients electing to undergo surgical intervention for propulsive metatarsalgia, whether wedge-cut osteotomy compared to flat-cut Weil (with or without adjunct procedures) is associated with increased pain relief during activities of daily living up to 12 months postoperatively. The secondary objective of this study is to determine whether a lower incidence of procedure-specific complications (floating toe or stiffness) at up to 12 months follow-up is associated with the wedge-cut procedure, compared to the flat-cut Weil.

## METHODS

### Design and setting of the study

The proposed study is designed as a randomised controlled superiority trial with two parallel groups comprising 1:1 allocation (Figure 1). Patient recruitment and data collection will be performed at the private hospitals and consulting clinics for the participating surgeons (New South Wales, Australia). Patients will be randomly allocated to either intervention using a central computer-based allocation randomisation system prior to surgery. The trial will be embedded in a clinical registry with patient, surgical and clinical data captured as part of routine clinical care.

**Figure 1:**
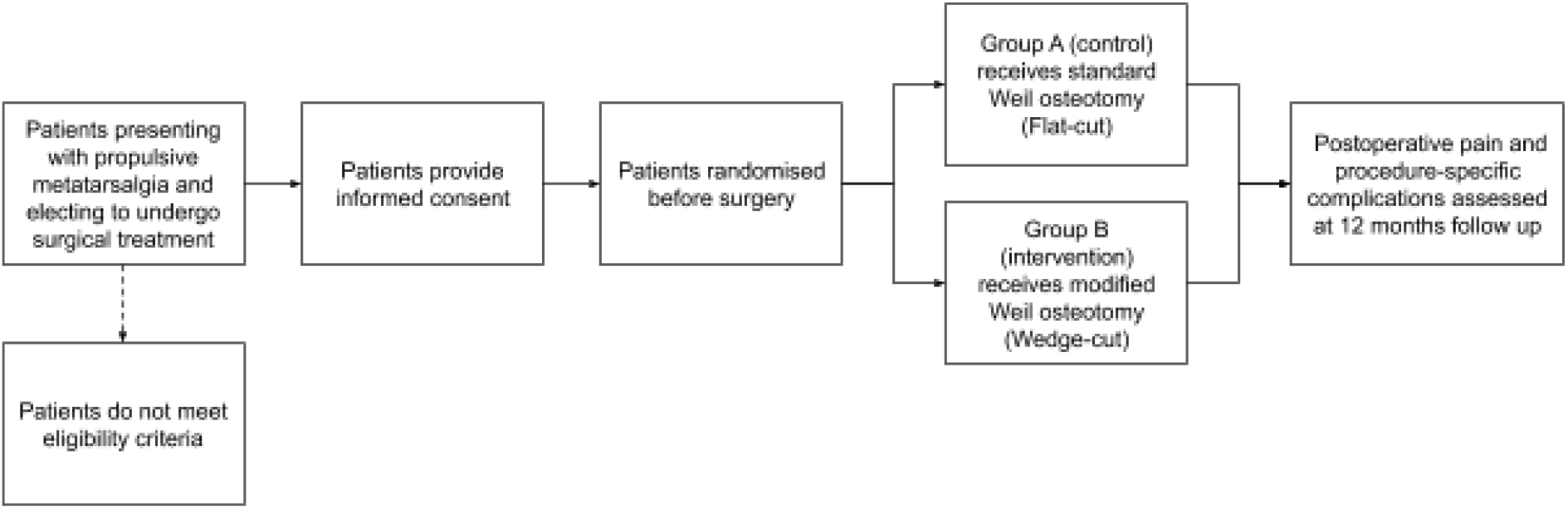
Study design for the randomised control trial.

The trial is scheduled to commence in January 2021 and due to be completed by June 2023. The study has been registered on an online registry for clinical trials (Australia New Zealand Clinical Trial Registry, ANZCTR), where study site and sponsor details are listed (ACTRN12620001251910). The study will be reported as per the CONSORT guidelines (Moher et al., 2010; Schulz et al., 2010).

### Data sources

Patient personal and medical data is routinely collected at the consulting clinics and entered into the practice management systems (Bluechip, MedicalDirector, Australia and Genie, Genie Solutions, Australia). The data is collated with patient reported outcome measures (PROMs) and organised into the Sydney Orthopaedic Foot and Ankle Research Institute (SOFARI) Registry established within the surgeon’s practice (ACTRN12620000331932), and will form the primary source for patient, clinical, intraoperative and postoperative data for the current trial. Ethical approval for use of the practice registry for research was provided by the New South Wales/Victoria branch of the Ramsay Health Care HREC (HREC approval number 2020-007). Registry data is hosted in a secure, HIPAA-compliant cloud-based database (Google Suite, Google, USA).

### Participants

#### Eligibility and Recruitment

Patients presenting to the participating surgeons for treatment of propulsive metatarsalgia who are over 18 years of age and eligible for surgical intervention (having failed a minimum of six months of conservative interventions) will be eligible for recruitment. Patients will need to be registered in the SOFARI registry and provide additional written informed consent for participation in the present randomised controlled trial.

Patients who have declined or revoked consent for use of clinical data for research (for the SOFARI registry or the current trial), or are unable to provide informed consent will be excluded. Patients will additionally be excluded if they have had recent (<6 months) prior surgery to the affected forefoot, if they require additional procedures involving the soft tissues of the foot-ankle complex, have had their surgery booking cancelled with the participating surgeon, or have been judged by the participating surgeons as incapable to complete PROMs as required for the study due to psychological impairment or insufficient English language capacity (Figure 2).

**Figure 2:**
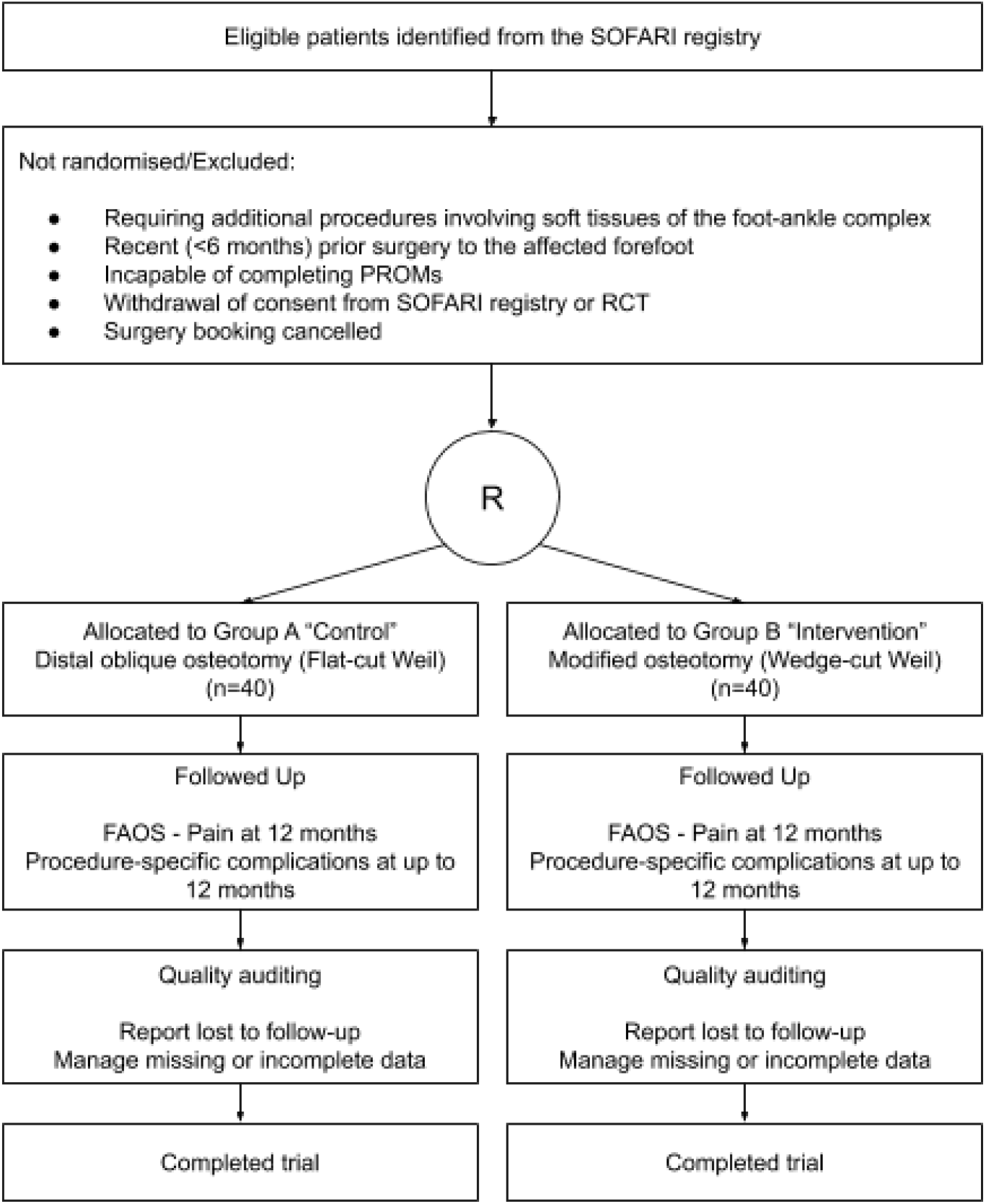
CONSORTdiagram (Schulz et al., 2010) with the key stages identifying patients that will be included in the analysis. *R* indicates randomisation.

### Interventions

#### Surgical technique

Eligible patients will be randomised in equal proportions between those receiving the flat-cut Weil osteotomy (the “control” intervention, Group A) and the wedge-cut Weil osteotomy (the “experimental” intervention, Group B).

Group A will receive an active control intervention. Preoperative planning will be undertaken to determine the amount of metatarsal shortening required to restore the parabola of the affected metatarsal. The amount of shortening will be determined by recreating the Maestro criteria for metatarsal parabola (Pascual Huerta et al., 2017). At the time of surgery, the patient will be prepared in the standard fashion, placed supine on the operating table with the operated limb prepared and draped to create a sterile area. General intravenous sedation will be administered in addition to prophylactic antibiotics and an ipsilateral ankle tourniquet is applied and inflated prior to the first incision. A semi elliptical transverse incision will be used to expose each metatarsophalangeal joint to be operated, spanning between the tendons of the extensor muscles without cutting or lengthening tendons. Additional care will be taken to avoid release of other soft tissues such the medial and lateral collateral ligaments.

The metatarsal and phalangeal joint surfaces will be inspected and additional resection performed to remove plantar capsule adhesions where required (DeCarbo & Dial, 2014). The metatarsal bone will be dissected, retracting other soft tissues, including the joint capsule to aid in visualisation and access to the metatarsal neck. A rasp will be used to remove the soft tissue on either side of the osteotomy location. The dorsal zone of cartilage inferior to the metatarsal head will be identified and used as a landmark for the osteotomy. Neurovascular bundles in the intertarsal space will be retracted and a microsagittal motorised saw held parallel to the weightbearing surface of the foot moved from the dorsal to plantar cortices of the metatarsal to dissect the head from the neck in preparation for repositioning. The plantar plate will be inspected and repaired as required and the metatarsal head shifted proximally to achieve the desired shortening, avoiding medial or lateral shifting.

The osteotomy will be stabilised with a Kirschner wire placed across the osteotomy (along the metatarsal shaft) to guide screw fixation. Once the screw is fully engaged the K-wire will be removed. The overhanging bone ledge will be resected with a bone saw or rasp, followed by releasing of the soft tissues under retraction closure of the joint capsule. The remaining soft tissues will be closed in layers and dressed with strips and bandages. The patient will be moved to recovery and discharged as per normal.

The preparation of the patient and surgery site for Group B will be as described for Group A (the control intervention). The surgical procedure will be modified by adding a second osteotomy proximally to remove a rectangular wedge of bone for the purposes of shortening the metatarsal and repositioning the floating segment of metatarsal head proximally to the metatarsal neck (Melamed et al., 2002).

Postoperative management for both groups will include elevation of the foot for the initial 72 hours after surgery, and stitches removed in 10-14 days. Analgesics and antibiotics will be prescribed for the first 14 days after surgery, with weightbearing as tolerated.

#### Modifications and adherence

If the participant requests a particular intervention, they will be unenrolled from the trial and their data will not be included in later analyses. Adherence to the randomised intervention allocation will be assessed by comparing the patient osteotomy group allocation as identified in the operation report, to the trial master sheet with the allocation information. Follow-up and PROMs compliance will be encouraged through follow-up reminders, as part of the SOFARI registry processes. The primary outcome will be collected electronically, with controls in place to prevent partial completion of the questionnaire.

#### Concomitant care

If required, the osteotomies (both control and interventional) are performed with adjunct procedures for toe correction, based on the individual case and patient’s anatomy:

- Proximal interphalangeal arthrodesis, which involves the longitudinal insertion of a Kischner wire or pin to fuse the most proximal joint of the lesser toe (Jay et al., 2016)
- Fusion/arthrodesis of the first MTP joint, which involves the longitudinal insertion of a Kirschner wire or pin to fuse the most proximal joint of the first ray (Donegan & Blume, 2017)
- Hallux valgus correction, which is a correcting osteotomy performed to realign the first ray in presence of first tarsometatarsal joint hypermobility (Bia et al., 2018)

### Outcomes

#### Primary outcome measures

The primary outcome of this trial will be the Foot and Ankle Outcome Score (FAOS) - Pain subscale, collected at up to 12 monthly postoperatively. The FAOS was developed as a foot and ankle-specific PROMs assessment analogous to the Knee injury and Osteoarthritis Outcome Score (KOOS), with content validity confirmed with 213 patients with ankle instability (Roos et al., 2001). The FAOS consists of 5 subscales; Pain, other Symptoms, Function in daily living (ADL), Function in sport and recreation (Sport(Rec), as well as foot and ankle-related Quality of Life (QOL). Patients rate the questions from on a 5-point Likert scale, with 0 representing no symptoms/problems, and 4 representing extreme symptoms. The FAOS has been used in patients with lateral ankle instability, achilles tendonitis and plantar fasciitis, with reliability confirmed in instability patients (Roos et al., 2001) and responsiveness confirmed in patients with achilles tendinosis (Roos et al., 2004) and hallux rigidus (Hogan et al., 2016). Given the interventions performed in this trial are to treat metatarsalgia, a condition characterised by pain during weight bearing activity, particularly noticeable during the push-off phase of the gait cycle, the pain subscale of the FAOS questionnaire was selected as the most clinically relevant patient-reported measure for the purposes of measuring the primary outcome of this trial. One study reported an MCID of 15.3 points (95% confidence interval 10-20.6) (Sierevelt et al., 2016), with a difference between groups equal to half this (7.7) deemed sufficient to guide future decision-making with respect to technique selection. For analysis purposes a mixed-effects model will be used, which will aggregate scores from all responders and report mean and standard deviation (SD) associated with different combinations of model predictors, such as intervention group, sex and adjunct procedures.

#### Secondary outcome measures

The secondary outcome of this trial will be the incidence of procedure-specific complications at up to 12 months postoperatively. The most commonly reported complication of the Weil osteotomy is a floating toe, with an overall occurrence of 36% (Highlander et al., 2011). A floating toe is defined as a toe that is not in contact with the floor under weight-bearing conditions (Migues et al., 2004). It will be recorded using a weight-bearing coronal view static photograph and manually rated. Recurrence is reported in 15% of the cases, followed by transfer metatarsalgia (7%) and delayed union, non-union, and malunion collectively reported in 3% of the cases (Highlander et al., 2011). Metatarsal osteotomy union will be defined using cortical continuity (DeSandis et al., 2015), with patients failing to show any healing (nonunion) and insufficient cortical continuity (delayed) by the 12 month review, identified in the trial data.

### Participant timeline

The time schedule for participant recruitment, interventions and assessment in this trial will follow the standard clinical pathway embedded within the SOFARI registry (Figure 2). The recruitment, allocation, surgery and 12 months follow-up time points relevant to the current trial are described in Table 1.

**Table 1:** Schedule of enrolment, interventions and assessment for the trial.

| TRIAL | STUDY PERIOD |  |  |  |  |
| --- | --- | --- | --- | --- | --- |
|  | Recruitment | Allocation | Surgery | Post surgery |  |
| Timepoint | -t<sub>2</sub> | -t<sub>1</sub> | 0 | Up to ~6 weeks | t<sub>1</sub> =12 months |
| **Enrolment** |  |  |  |  |  |
| Eligibility screen | X |  |  |  |  |
| Informed consent | X |  |  |  |  |
| Allocation |  | X |  |  |  |
| **Interventions** |  |  |  |  |  |
| Group A (control intervention) |  |  | X |  |  |
| Group B (experimental intervention) |  |  | X |  |  |
| **Assessments** |  |  |  |  |  |
| Patient demographic and clinical data | X |  |  |  |  |
| Intraoperative data |  |  | X |  |  |
| Serious adverse event assessment |  |  |  | X |  |
| FAOS - Pain |  |  |  |  | X |
| Complications data |  |  |  |  | X |

### Sample size

The sample size required for the primary outcome was established to provide adequate power to detect half an MCID difference (15.3/2) in the FAOS Pain score between the control flat-cut Weil and experimental wedge-cut groups, with an average estimated baseline standard deviation of 20.8. The MCID and baseline SD were derived from a previous study (Sierevelt et al., 2016), and 0.5MCID was determined to be a clinically important effect that would influence future clinical decision-making regarding technique selection. A mixed-effects model (analysis of covariance, ANCOVA) was selected as the most appropriate to answer the question posed, with power (□) at 0.9 and α of 0.05, with an allowance for a dropout rate of 10%. The required sample size was determined using GPower (Faul et al., 2007) to be N=80 patients, with patient age at surgery, sex, body mass index, single/multiple procedures and adjunct procedures included as model covariates.

The sample size required for the secondary outcome was established to provide adequate power to detect a 15% reduction in complication incidence from a baseline of 60% incidence. The reduction in incidence was deemed to be the minimal amount of improvement that would influence future decision-making regarding technique selection in this patient population. A one-sided test within a logistic regression model was selected within GPower, with the R^2^ for other predictors in the model (patient age at surgery, sex, body mass index, single/multiple procedures and adjunct procedures) set to 0.1. The required sample size was determined to be N=123 procedures.

### Recruitment

The Principal Investigator for the study performed a total of 127 Weil osteotomies in 87 patients over the last 12 months. To meet the required sample size of N=80 patients with N=123 procedures to assess the primary and secondary outcomes respectively, the expected recruitment period will extend over 12 months. All prospective patients eligible for recruitment will be approached to participate in the study.

### Allocation

Participants will be randomly assigned to receive either a flat-cut Weil osteotomy (control, Group A) or wedge-cut osteotomy (experimental, Group B) procedure using a central computer-based simple randomisation system, with a 1:1 allocation without blocking. The random allocation sequence will be embedded within the clinical registry via a randomisation algorithm (Matlab 2018b, Mathworks Inc, USA). The allocation will be communicated electronically to the treating surgeon via an allocation code comprising nondescript terminology, whereby the allocation cannot be inferred from the label to other study personnel. The system will not release the allocation code to the treating surgeon until the patient has been recruited into the trial to maintain allocation concealment.

### Blinding

Blinding is not feasible for the treating surgeon or the senior data engineer responsible for coding the randomisation sequence, who will have access to the allocation code and allocation code key. The senior data engineer will not have further involvement in the study beyond the allocation and data coding.

Trial participants will be blinded to the intervention assignment, and will be counselled at the time of recruitment regarding both surgical techniques, supplementing the information provided on the Patient Information Sheet and Consent Form (PISCF). Surgical assistants, practice staff, clinical outcome assessors members of the research team and study statistician involved in the data analysis will be blinded to the allocation, with access to only the allocation code which will comprise a label where grouping is not able to be inferred.

Postoperative clinical evaluations and assessment of serious adverse events will be conducted by a surgical fellow that is hosted within the clinic on a rotating basis of six months. In cases where the surgical fellow assisting with surgery also performs the postoperative evaluation, masking will occur through access to only the allocation coding described above. While the treatment evaluator will have access to postoperative radiographs of the operated structures within the foot, it is expected the intervention allocation cannot be derived from these images. Patients returning for evaluation at the 12 month follow-up will be assessed by a fellow not involved in the surgery.

#### Unmasking

In situations where an adverse event or complication has occurred such that reoperation or other invasive intervention is deemed necessary, it may be necessary to unmask the intervention allocation to plan appropriate treatment. Requests to unmask will be made electronically via the treating surgeon and logged within the study masterlist. Requests will be managed by the senior engineer with access to the allocation key.

## DATA COLLECTION METHODS

### Data collection

Patient personal and medical data is routinely collected and stored in the participating surgeons’ practice management systems (Bluechip, MedicalDirector, Australia and Genie, Genie Solutions, Australia). Patient demographic data, comorbidities and full patient history relating to the history and onset of the foot and ankle condition will be recorded in the practice management system during patient consultation. Radiological reports collected routinely as part of diagnosis, surgical planning and postoperative follow-up, full description of diagnosis, mode of treatment and details of nonoperative and surgical interventions, and timing of treatments will also be recorded in the practice management system during patient consultation. Intraoperative data, patient-reported outcomes data and findings from clinical examination will be electronically entered into the SOFARI registry via web-based forms (Google Suite, Google, USA) by the clinical research nurse, research team, surgeon or by the patient.

Intraoperative (surgical technique), patient-reported (FAOS-Pain) and clinical outcomes (complications) data collected during this trial will additionally be linked to the SOFARI registry via the same modes of data entry. Data collection will continue until the minimum sample size requirements for the study are met with complete records established.

#### Participant retention

Once a participant is enrolled in the study, they will be contacted at 3, 6 and 12 months postoperatively for PROMs follow-up data collection as per the existing SOFARI registry procedures. The routine contact will facilitate participant retention up until the 12 month follow-up. Follow-up reminders will be administered from the clinic, and the electronic questionnaires can be completed remotely at the patient’s convenience. There is an additional opportunity to collect PROMs data when the patient visits the clinic for their 12 month follow-up, if they have not completed the electronic forms.

In cases where the intervention protocol is not followed, the participant will be removed from the trial and the randomisation slot filled by the next participant recruited into the trial.

### Data management

#### Data entry

All data collected in this trial will be entered electronically via web-based forms (Google Suite, Google, USA) by either the investigators, research team or participants. The research team will be responsible for organisation of the data within a HIPAA compliant database environment (Google Suite, Google, USA) and performing quality assurance checks on the consolidated dataset.

#### Clinic data management

Identifiable data (collected only as necessary for treatment and management of the health services) will be stored permanently on the practice management software, as is standard clinical practice. The data will be stored on a secure server within the consulting rooms, with restricted access.

#### Data linkage processes

Identifiable personal and medical information collected from patients will be stored and kept indefinitely within the database to link patient records to patient information from other sources (e.g. electronic medical records within the practice management systems, clinic notes or radiology records). The research team will access the practice management software in order to conduct quality assurance audits which will involve access to identifiable data, within the environment of the password protected server.

Web-based forms will be provided to patients to capture validated PROMs using identifiers to link the form responses to the rest of the registry dataset. No identifiable information (name, date of birth, address, contact information) will be included or requested of patients in the forms. Standard operating procedures for data entry will be available to individuals who will require access to the database via a web browser interface to streamline and control data-entry processes. All members with access to the data will have signed a non-disclosure agreement.

Access to the trial data will be password protected. Data will be stored in a HIPAA compliant environment (Google Suite, Google, USA) prior to further processing locally by the research team. The research team will retain access to the data for the duration of the trial. At the termination of the trial, any study-relevant data will be transferred to the clinic servers and deleted from the research team’s environment. The data gathered will be retained on the existing password protected servers of the clinic for future reference, publications and potential future studies.

Supplementary information within clinical notes in external databases, such as those at the hospitals where the treatment or surgery is performed or radiology services, will not be integrated directly with the study database. This data will be requested from these systems on an as-needed basis by the surgeons or practice managers, matching patients by name, date of birth and treatment, and will be manually transcribed into the practice management system and subsequently into the database.

## STATISTICAL METHODS

### Principles

An analyst blinded to treatment allocation will perform the primary and secondary analyses, by comparing the intervention group to the control group. Data will be analysed following intention-to-treat principles (i.e. participants will be analysed in the group to which they were randomised to). Data normality of baseline characteristics and process measures will be evaluated by visual inspection of histograms. Continuous variables will be presented as means and standard deviations for normally distributed variables: median, minimum, maximum, and interquartile range for non-normally distributed variables. Frequencies and percentages will be used to summarise categorical variables. Percentages will be calculated using the number of participants for whom data are available as the denominator. Alpha will be set at 0.05 and effect sizes as described in the sample size calculation will be considered of interest.

### Data integrity

All data points for the study core dataset will be retrieved electronically, so comparing to original paper records will not be necessary. Data integrity will be defined in the context of this study as completeness, consistency and validity. An error rate of <3% will be considered acceptable across the total of fields assessed.

Patients who are otherwise lost to follow-up will be included in the analysis under an intention-to-treat framework. Where appropriate, imputation will be used to provide estimates of outcomes for patients lost to followup. To mitigate the effects of loss to follow up on the analysis, the sample size factored in an estimated dropout rate of 10% to ensure adequate power for analyses requiring listwise deletion of missing data.

### Evaluation of demographics and baseline characteristics

Baseline characteristics will be presented in a table stratified by treatment group. Hypothesis testing of baseline characteristics between groups will not be performed in accordance with the recommendations of the CONSORT statement (Schulz et al., 2010). Figure 3 describes the prognostic factors which will be treated as potential confounders of the effect of the intervention and included in the models used to analyse primary and secondary outcomes. These confounders were selected based on the available literature on pain ratings in metatarsalgia (Chou et al., 2008; Hurn et al., 2014; Khan et al., 2011; Lopez & Slullitel, 2019; Park et al., 2016).

**Figure 3.**
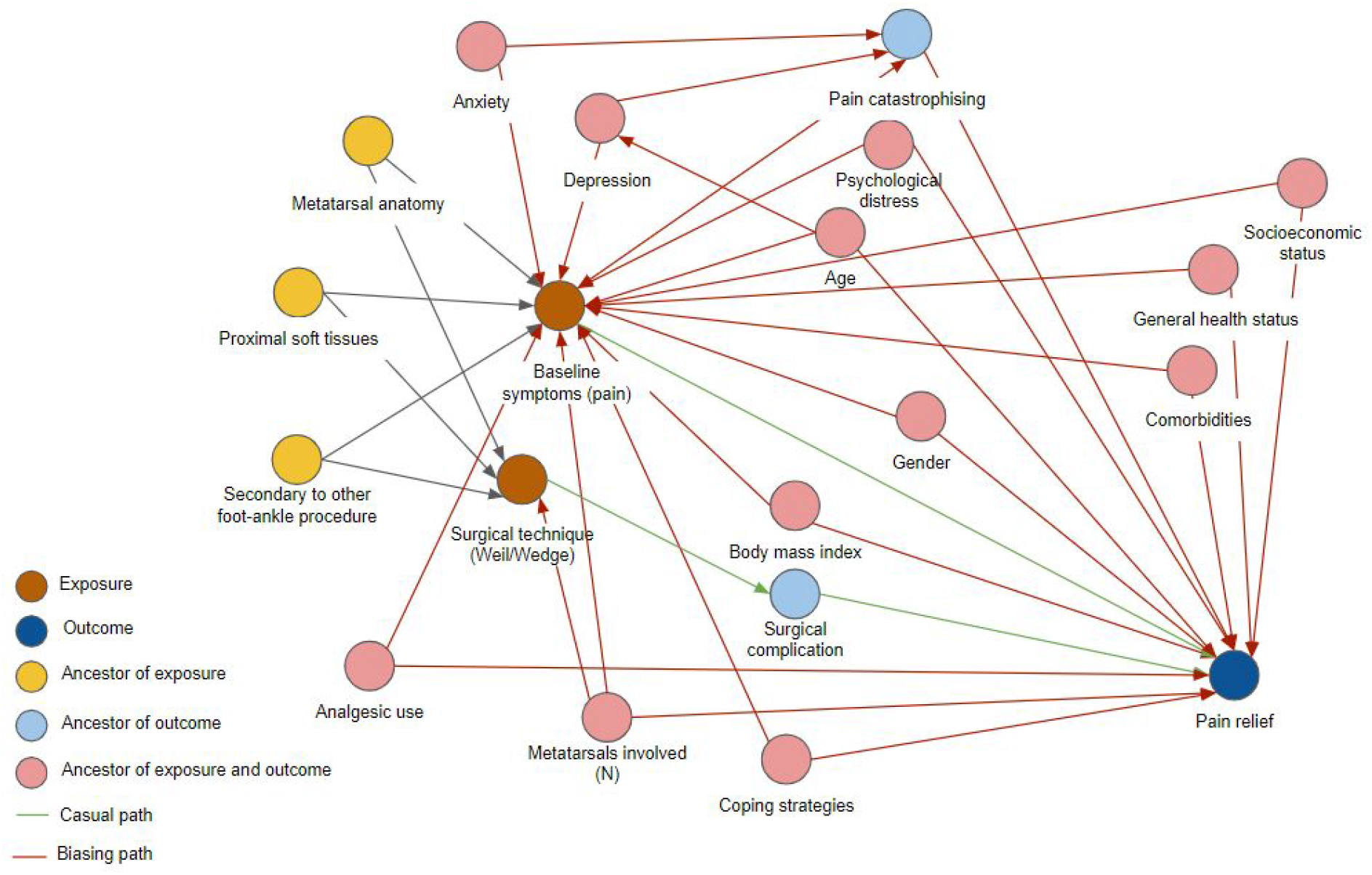
Directed acyclic graph (DAG) or concept map indicating potential confounders for analysis of the study outcomes.

### Primary Outcome Analysis

The primary outcome of the trial is postoperative pain at 12 months follow-up represented by the pain subscale of the FAOS. A mixed-effects ANCOVA will be used to test for differences between groups with the following factors and covariates included in the model:

- Patient ID (random factors)
- Baseline FAOS pain subscore (covariate)
- Age at surgery (covariate)
- Sex (covariate)
- Multiple procedures
- Adjunct procedures

The overall model fit will be assessed with partial eta^2^ and coefficients (□) with 95% confidence limits will describe the strength and direction of relationships between factors and the postoperative FAOS-Pain score. Post-hoc comparisons with Dunnett tests (against control) will be used to compare osteotomy groups. Cohen’s d will be used to report effect size of differences between groups. A Cohen’s d exceeding 0.36 will be deemed clinically significant.

### Secondary Outcome Analysis

The secondary outcome of the trial will be the incidence of procedure-specific complications by the end of the follow-up period (12months). A binary logistic regression will be used to assess the effect of intervention allocation on complications incidence in a multivariable model containing confounders described above. Alpha will be set at 0.05 and partial eta^2^ used to assess model fit. Odds ratios with 95% confidence limits will describe the strength and direction of relationships between model predictors and the probability of having a complication compared to not having one.

### Additional analyses

Depending on the overall incidence of procedure-specific complications (assuming that the incidence is >5% overall) a Cox regression will be employed to compare groups for time to event (complication detected) between day 1 postoperatively to 12 month evaluation with right censoring for participants that complete the study follow-up with no complications reported. Hazard ratios with 95% confidence intervals will be used to describe the strength and direction of the relationship between intervention allocation and time to event.

### Analysis population

The nature of the dataset does not lend itself to multiple imputation in the first instance. If patients have not returned the form at follow-up, despite multiple reminders, a multiple imputation approach will be employed based on published guidelines for analysis of trial data (Jakobsen et al., 2017). Secondary outcomes are unlikely to have missing data due to the nature of clinical follow-up and the acute nature of procedure-specific complications (i.e. all patients are reviewed in person or by phone within the timeframe that complications would be observed). In the event that missing secondary outcomes are apparent, analyses will be performed to assess the *randomness* of the missing data and to determine whether complete case analysis is appropriate. In the event that the missing data does not conform to a random pattern, sensitivity analysis using worst-case, best-case scenarios for missing data will be constructed and reported (Jakobsen et al., 2017).

## MONITORING

### Data monitoring

A governance and steering committee with the Principal Investigators and the research team will maintain the governance of the trial. The scope of the proposed protocol does not warrant a data monitoring committee due to the nature of the interventions and the outcomes selected for comparison between groups.

### Interim analyses

No interim analysis has been planned that would lead to early termination based on the results of the study. The study will be monitored for adverse events on a continual basis and reported regularly to the investigators, and an unacceptably high incidence in either treatment group (50% greater than reported in the equivalent literature) may be reviewed by the investigators and cause the trial to be terminated.

### Harms

An adverse event is defined as any deviation from the normal recovery trajectory requiring alteration to the patient’s care or medical intervention. Adverse events will be collected after the subject has provided consent and enrolled in the trial. A serious adverse event (SAE) for this study is any untoward medical occurrence within the follow-up period of the study and results in any of the following: Life-threatening condition, severe or permanent disability, prolonged hospitalisation, or a significant hazard. Investigators will determine relatedness of an event to the trial based on a temporal relationship to the intervention, as well as whether the event is unexpected or unexplained given the subject’s clinical course, previous medical conditions, and concomitant medications. A separate form will be set up for adverse reporting, to be completed by the evaluator or the trial monitoring team based on updated clinical notes for the patient within the practice management system and electronically linked to the study database.

### Auditing

Quality assurance of the trial will be maintained through auditing and reporting of data completeness, consistency and validity: completeness (all data fields required are filled in) will be monitored in real time and compared to the requirements outlined in the core dataset; consistency (data responses match the rules specified within the core dataset) will be examined by assessing continuous variables for outliers and categorical variables for consistency with pre-specified responses within the core dataset; validity (data is accurate and correct) will be assessed in a subsample of patients (10%) by spotchecking the information held in the study masterlist relative to the original source in the practice management system. Discrepancies will be investigated and rectified (where possible) by practice staff or the research team. Audit reporting will be provided to the registry steering committee and communicated to the Principal Investigators on a quarterly basis.

## ETHICS AND DISSEMINATION

### Ethical approval and protocol amendments

Ethics approval for this study was provided through the NSW/VIC branch of the Ramsay Health Care Human Research Ethics Committee (HREC approval number 2020-007). Any modifications to the protocol which may impact on the conduct of the study, patient safety, or significant administrative aspects (e.g. changes to study sponsorship) will require a formal amendment to the protocol. Such amendments will be agreed upon by the investigators and approved by the NSW/VIC Ramsay Health HREC prior to implementation. Modifications will also be reflected in the public trial registry record on the ANZCTR.

### Consent or assent

Participants will be prospectively recruited at their initial clinic for the trial. All potential participants will receive a Participant Information and Consent Form (PISCF) which will outline the information that will be collected for the study, how that information will be used, with whom it will be shared, and how to opt-in to the study if they so wish. The information sheet will also inform the patient of the voluntary nature of the research, and that they may withdraw their consent and information from the study at any time, without impacting their treatment.

If a patient chooses not to opt in, clinic staff will inform the research team and note this in their practice management system and the study database so patients are not approached. Patients who wish to cease participation in the study will have their status on the study database marked as “Withdrawn from trial” and their information will not be used in any subsequent research or analyses of the data.

Patients who do not have an acceptable level of English will be asked to complete medical forms and questionnaires if they are accompanied by a translator of an acceptable standard as determined by the treating surgeon. The standard must be such that the patient is able to complete the required forms/questionnaires and understand their obligations and rights.

### Ancillary studies

Potential uses of the datasets generated and/or analysed during the trial may include subsequent analysis of subgroups for investigation of surgical/management and patient centred outcomes for internal clinic purposes.

The research data may form the basis of potential future studies, including additional research sites within allied health networks or collaborations with other research groups for biomechanical evaluation of patients. Where appropriate, open-access principles will be followed to enable collaborative works with aligned groups, where the interest is academic and not for commercial uses, under appropriate licensing arrangements of deidentified data. Any future studies requesting use of this data will seek appropriate amendment of the relevant ethical approval.

### Confidentiality and data access

Confidentiality and privacy of patient information in the trial will be protected through the execution of nondisclosure agreements between all involved parties, prohibiting them from sharing identifiable information externally.

Identifiable trial data is kept electronically on a secure server at the study clinic sites, and de-identified study data will be stored in a secure, HIPAA-compliant online database with access restricted to individual staff members responsible for handling the data.

No identifiable data from the study will be externally shared without consent from the patients. All identifying information such as name, date of birth, email or phone will be removed from any data prior to transfer of this data to sites not listed in the document. Patient identification numbers will be used as a substitute to identifiable data, and only the investigators will be able to re-identify patients. Patients will not be identified in any publication or presentation resulting from the trial.

All investigators will have unrestricted access to the cleaned data sets. Study data sets will be housed and secured as per the data management procedures outlined in this protocol.

### Ancillary and post-trial care

Patients that are enrolled into the study are covered by indemnity for negligent harm through standard medical indemnity insurance. The intervention is routine and well-practiced, and the insurance covers non-negligent harm associated with the protocol. This includes cover for additional health care and compensation or damages, whether awarded voluntarily or by claims pursued through the courts. Incidences judged to arise from negligence (including those due to major protocol violations) will not be covered by study insurance policies.

### Dissemination policy

#### Trial results

There are no restrictions in the public dissemination of results within the current scope of this protocol. Results will primarily be communicated via peer-reviewed publications and abstracts. Planned publications include a manuscript summarising findings from the trial to be published at the conclusion of the study. Patients will not be identified in any publication or presentation that will be published as a result of the trial. Patients are able to request the results of the trial and any resulting publications by contacting the clinic.

The results from the study may also be informally shared by the Investigators with peer networks in internal meetings, and be made available upon request to the ethics board and other health/regulatory authorities.

#### Authorship

For any resulting publications, authorship eligibility for anyone involved in the study design, management and conduct of the trial will be determined in accordance with the International Committee of Medical Journal Editors (ICMJE) authorship guidelines (ICMJE, 2019). Anyone not meeting the requirements for authorship (minimum 10% contribution) will be listed in the acknowledgements section of the manuscript.

#### Reproducible research

Supplementing this protocol manuscript, the statistical code used in the trial will be uploaded to GitHub (GitHub Inc, USA) to enable peer review where appropriate. The deidentified study dataset will be made available upon request, keeping in line with open access principles to enable collaboration, where the interest is academic and not for commercial uses. Only deidentified data will be shared if required, under the appropriate licencing arrangements.

## DISCUSSION

The trial will provide clinical data pertaining to the efficacy of the modified wedge-cut Weil osteotomy procedure compared to the traditional flat-cut method. Flat-cut Weil osteotomy is a routinely performed procedure with complications frequently reported (Besse, 2017; Highlander et al., 2011; Lunz et al., 2010). The wedge-cut modification purportedly has functional and mechanical advantages over the flat-cut technique, however in-vivo data and quality of evidence is currently lacking (Garg et al., 2008; Melamed et al., 2002).

A previous study reported satisfactory performance on the American Orthopaedic Foot and Ankle Society (AOFAS) forefoot score (with 65% of patients reporting no pain and 23% reporting mild pain), but higher than expected complication rates for transfer metatarsalgia, floating toes, infection, and wound healing complications in patients receiving segmental resection (wedge-cut) osteotomies (Garg et al., 2008; Melamed et al., 2002). The study however, was a retrospective case series, lacking baseline data and with a short (minimum 6 months) follow-up. To the best of the authors knowledge, the current trial will be the first to directly examine the clinical efficacy of the wedge-cut Weil osteotomy within a randomised control design.

Quality assurance of the trial will be maintained through regular auditing and reporting on data completeness, consistency and validity. The trial has been designed in accordance with the Standard Protocol Items: Recommendations for Interventional Trials (SPIRIT) 2013 guidelines, in order to enhance transparency and facilitate appraisal of its scientific merit, ethical considerations, and safety aspects (Chan et al., 2013).

## Supporting information

SPIRIT Checklist

Patient Information Statement and Consent Form

## Data Availability

N/A

## Author contributions

AW, MS and CS contributed to the conceptualisation of the study. CS, MF and NE contributed to the design of the study. MF and CS contributed to drafting the study protocol, with critical feedback provided by NE and clinical guidance provided by MS and AW. AW sponsored the study.

## Acknowledgements

The authors would like to acknowledge Meredith Harrison-Brown for her assistance with study planning and manuscript administration, and Milad Ebrahimi for this assistance with study design.

