## Supplementary material for "The effect of osteotomy technique (flat-cut vs wedge-cut Weil) on pain relief and complication incidence following surgical treatment for patients presenting with metatarsalgia in a private metropolitan clinic: Protocol for a randomised controlled trial": Patient Information Statement and Consent Form

### Participant Information Sheet/Consent Form

| | |
|---|---|
| **Title** | The effect of osteotomy technique (Weil vs modified Weil "Wedge") on pain relief and complication incidence following surgical treatment for patients presenting with metatarsalgia |
| **Short Title** | The effect of osteotomy technique on pain relief and complication incidence of surgical treatment of metatarsalgia |
| **Project Sponsor** | Dr Andrew Wines |
| **Principal Investigator** | Dr Andrew Wines |
| **Associate Investigator(s)** | Dr Michael Symes; Dr Corey Scholes |
| **Locations** | North Sydney Orthopaedic and Sports Medicine Centre<br>Suite 2, Mater Clinic, 25 Rocklands Road<br>Wollstonecraft NSW 2065<br><br>Sydney Orthopaedic Trauma & Reconstructive Surgery<br>Suite 201, 131 Princess Highway<br>Kogarah NSW 2217 |

#### Part 1 What does my participation involve?

##### 1 Introduction

You are invited to take part in this research project, because you are enrolled in the surgeon's practice registry and require treatment for propulsive metatarsalgia. The research project is testing a new surgical method for the treatment of propulsive metatarsalgia, called a modified Weil "wedge" osteotomy.

This Participant Information Sheet/Consent Form (PISCF) tells you about the research project. It explains the tests and treatments involved. Knowing what is involved will help you decide if you want to take part in the research. Please read this information carefully. Ask questions about anything that you don't understand or want to know more about. Before deciding whether or not to take part, you might want to talk about it with a relative, friend or your local doctor.
Participation in this research is voluntary. If you don't wish to take part, you don't have to. You will receive the best possible care whether or not you take part.

If you decide you want to take part in the research project, you will be asked to sign the consent section. You will be given a copy of this PISCF to keep. By signing it you are telling us that you:

• Understand what you have read
• Consent to take part in the research project
• Consent to have the treatments that are described
• Consent to the use of your personal and health information as described.

##### 2 What is the purpose of this research?

Your surgeon routinely performs Weil osteotomies to treat propulsive metatarsalgia. Several variations of the Weil osteotomy procedure are available, but the two variations routinely performed by your surgeon are the flat-cut and wedge-cut techniques. During the flat-cut procedure, a single oblique cut is made on the affected metatarsal head, and the head is shifted back to shorten the bone, before it is fixed into position. In the wedge-cut technique , two cuts are made, enabling a "wedge" or small slice of the bone to be removed, in order to shorten the affected metatarsal. The wedge-cut technique is not new or experimental , however, preliminary evidence suggests that this technique is able to reduce complications that are typically associated with the traditional flat-cut technique by stabilising the joint and improving muscle function.

The clinical evidence supporting this is limited, and the two techniques have not previously been compared before. The proposed study has therefore been initiated by your surgeon to investigate whether the modified Weil osteotomy with removal of a “wedge” compared to the traditional Weil technique is associated with better pain relief and fewer complications at up to 12 months after surgery.

#### 3 What does participation in this research involve?

Your participation in this research will primarily involve the standard of care for surgical treatment of propulsive metatarsalgia. After the initial consultation, your surgeon will discuss whether you are an appropriate candidate for Weil osteotomy, and invite you to take part in this study. Your treatment will involve clinical consultations to understand your symptoms and clinical history, followed by elective surgery in hospital under the care of your surgeon. Written consent will be obtained from you prior to any study assessments being performed.

If you choose to participate, you will take part in a randomised controlled trial (RCT). RCTs are designed to compare different procedures by putting people into groups and giving each group a different treatment, in order to determine which treatment is best for treating a condition. The results are compared to see if one is better. To try to ensure the groups are the same, each participant is randomly allocated to a group.

In this study, you have a 50% chance of being allocated to the modified Weil “wedge” group, instead of the traditional Weil osteotomy. You will be blinded to the treatment you are receiving, but your surgeon will know which treatment you are receiving at the time of surgery. As part of the routine treatment schedule, your surgeon will assess you 12 months after surgery. The assessment will involve a consultation of any complications experienced after surgery, completion of a short pain questionnaire and a static photograph taken to assess the incidence of a floating toe.

This research project has been designed to make sure the researchers interpret the results in a fair and appropriate way and avoids study doctors or participants jumping to conclusions. There are no costs associated with participating in this research project, nor will you be paid. This research will not require any additional participation or travel outside what is generally expected from patients to attend their consultations and undergo surgery.

#### 4 What do I have to do?

Participation in this research project will not require any additional restrictions or modifications to your lifestyle, diet or medications, other than what has been advised by your surgeon as part of your routine treatment and care. You will be asked to attend the clinic for follow-up assessment and additional measurements, and be asked to fill out reporting tools to collect outcomes data.

#### 5 Do I have to take part in this research project?

Participation in any research project is voluntary. If you do not wish to take part, you do not have to. If you decide to take part and later change your mind, you are free to withdraw from the project at any stage. Your decision whether to take part or not to take part, or to take part and then withdraw, will not affect your routine treatment, your relationship with those treating you or your relationship with your surgeon. If you do decide to take part, you will be given this Participant Information and Consent Form to sign and you will be given a copy to keep.

#### 6 What are the alternatives to participation?

You do not have to take part in this research project to receive treatment by your surgeon. If you do not wish to participate in this study, you will not be randomly allocated to a specific group. Your surgeon will opt to perform either the Weil osteotomy or modified “wedge” procedure based on their clinical experience, as both techniques form a routine part of their surgeries. If you have concerns about your treatment options, you can also discuss the options with your treating surgeon.

**7 What are the possible benefits of taking part?**

There will be no clear benefit to you from your participation in this research. However, the study will assess the clinical efficacy of the modified Weil “wedge” procedure, and has the potential to offer improved treatment options for surgeons that may be of benefit to future patients.

**8 What are the possible risks and disadvantages of taking part?**

There are no differential risks between the two arms of the study for patients taking part in the study. However, your surgeon will discuss with you any possible risks generally associated with undergoing surgical treatment. These include: infection; problems with wound healing; nonunion of bone; nerve injury causing numbness; tingling and/or pins and needles; blood vessel injury; deep venous thrombosis/ pulmonary embolism (which increases with smoking, oral contraceptive pill, hormone replacement therapy, immobility and obesity); ongoing pain; anaesthetic complications; and drug allergies.

If you choose to participate in this study, you will be randomly allocated to one of two groups - those receiving the traditional Weil osteotomy, and those receiving the modified Weil “wedge” procedure. Both procedures are routinely performed by the surgeon, and are generally safe. Complications that may arise from the traditional Weil procedure include a floating toe (defined as a toe that is not in contact with the floor when standing, occurring in about 36% of cases), recurrence of metatarsalgia (15% of cases), transfer of metatarsalgia to the other toes (7%), and stiffness of the toe (incidence unknown). It is not known whether the modified Weil “wedge” procedure will result in equivalent, better or worse outcomes, which is why this study has been designed to investigate this. While it is not expected that you will experience any serious side effects, tell your doctor immediately about any new or unusual symptoms that you get. This applies regardless of whether or not you choose to participate in the study.

Due to the current COVID-19 global pandemic, the clinic has also implemented infection risk minimisation practices in line with the national and state health and safety guidelines. These practices will be adhered to regardless of your decision to participate in the study.

There is a very small risk to your privacy through data loss or a leak from the electronic storage. This risk has been minimised by using industry-standard security safeguards and restricting access to a few trusted individuals.

**9 What if new information arises during this research project?**

Sometimes during the course of a research project, new information becomes available about the treatment that is being studied. If this happens, your surgeon will tell you about it and discuss with you whether you want to continue in the research project. If you decide to withdraw, your surgeon will make arrangements for your regular health care to continue. If you decide to continue in the research project you will be asked to sign an updated consent form.

Also, on receiving new information, your surgeon might consider it to be in your best interests to withdraw you from the research project. If this happens, they will explain the reasons and arrange for your regular health care to continue.

**10 Can I have other treatments during this research project?**

As part of your routine consultation and treatment, your surgeon will explain to you whether any modifications or restrictions need to be applied to your lifestyle, diet or medications you are taking as a result of your treatment and surgery. Participating in this study will not affect or change the nature of this advice.

**11 What if I withdraw from this research project?**

If you decide to withdraw from this research project, please notify the practice manager or surgeon during your clinic visit, or via phone or email. Your status on the study database will be marked as “Withdrawn from research”, and your information will not be used in any subsequent research activities. If you do withdraw your consent during the research project, it will not impact the standard of care you receive from your surgeon or the clinical team.

##### 12 Could this research project be stopped unexpectedly?

Sometimes a research project may be stopped unexpectedly due to unforeseen circumstances, such as a high incidence of adverse events. While this is unlikely to occur, the study will be monitored for adverse events. An unacceptably high incidence in either treatment group may be reviewed by the investigators and cause the trial to be terminated. No interim analysis has been planned that would lead to early termination based on outcomes data alone.

##### 13 What happens when the research project ends?

At the conclusion of the study, you will be able to request the results of the study and any resulting publications by contacting the clinic if you wish. Findings from this research will be published in peer-reviewed scientific journals and presented at national and international conferences. Participants will not be identified in any publication, and reports and publications will be made available on request to the ethics board and other health/regulatory authorities.

#### Part 2 How is the research project being conducted?

##### 14 What will happen to information about me?

The research will involve the collection of clinical, demographic and surgical data by the clinic or your surgeon before, during and after surgery. By signing the consent form, you consent to your surgeon and relevant research staff collecting and using personal information about you for the research project. This data will be entered into a database to collectively compare the traditional and modified Weil procedures for all patients who have undergone surgical treatment of propulsive metatarsalgia under the care of your surgeon. The study will be conducted over three years, in order to recruit the required number of participants.

The data collected from you will be individually identifiable (via your name and surgery date), but will be converted to a re-identifiable (coded) format for the purpose of the analysis, with the use of treatment and patient identification numbers. Identifiable data will be used for data linkage purposes only. Research data will be stored in a database with a password-protected user interface, and will be retained exclusively by the researchers for a minimum of five years following the cessation of this research. EBM Analytics will have access to the research data for the duration of their engagement as research managers, but will no longer have access once this is complete.

It is anticipated that the results of this research project will be published and/or presented in a variety of forums. In any publication and/or presentation, information will be provided in such a way that you cannot be identified, except with your permission. In accordance with relevant Australian and/or NSW privacy and other relevant laws, you have the right to request access to the information collected and stored by the research team about you. You also have the right to request that any information with which you disagree be corrected. Please contact the research team member named at the end of this document if you would like to access your information.

Any information obtained for the purpose of this research project that can identify you will be treated as confidential and securely stored. It will be disclosed only with your permission, or as required by law. Only de-identified data may be made available from the corresponding author upon reasonable request. Potential future uses of the data obtained during this study may include subsequent analysis of subgroups for investigation of surgical management and patient centred outcomes for internal clinic purposes.

##### 15 Who is organising and funding the research?

This research project is being conducted by your surgeon, with the support of EBM Analytics. EBM Analytics is an independent research organisation engaged by the sponsor to assist with the research and perform data analysis.

You will not benefit financially from your involvement in this research project even if, for example, the knowledge acquired from analysis of your data proves to be of commercial value to the study sponsor. No member of the research team will receive a personal financial benefit from your involvement in this research project (other than their ordinary wages).

#### 16 Who has reviewed the research project?

All research in Australia involving humans is reviewed by an independent group of people called a Human Research Ethics Committee (HREC). The ethical aspects of this research project have been approved by the HREC of Ramsay Health Care .

This project will be carried out according to the National Statement on Ethical Conduct in Human Research (2007). This statement has been developed to protect the interests of people who agree to participate in human research studies.

#### 17 Further information and who to contact

If you want any further information concerning this project or if you have any medical problems which may be related to your involvement in the project (for example, any side effects), you can contact your surgeon’s practice manager.

**Clinical contact person**

| | |
|---|---|
| Name | Frances Sellars |
| Position | Practice manager/Secretary to Dr Wines |
| Telephone | (02) 9409 0563 |
| Email | |

If you have any complaints about any aspect of the project, the way it is being conducted or any questions about being a research participant in general, then you may contact:

| | |
|---|---|
| Reviewing HREC name | Ramsay Health Care |
| HREC Executive Officer | Professor Phil Mitchell |
| Telephone | (02) 9433 3854 |
| Email | |

### Consent Form

| | |
|---|---|
| **Title** | The effect of osteotomy technique (Weil vs modified Weil "Wedge") on pain relief and complication incidence following surgical treatment for patients presenting with metatarsalgia |
| **Short Title** | The effect of osteotomy technique on pain relief and complication incidence of surgical treatment of metatarsalgia |
| **Project Sponsor** | Dr Andrew Wines |
| **Principal Investigator** | Dr Andrew Wines |
| **Associate Investigator(s)** | Dr Michael Symes; Dr Corey Scholes |
| **Locations** | North Sydney Orthopaedic and Sports Medicine Centre<br>Suite 2, Mater Clinic, 25 Rocklands Road<br>Wollstonecraft NSW 2065<br><br>Sydney Orthopaedic Trauma & Reconstructive Surgery<br>Suite 201, 131 Princess Highway<br>Kogarah NSW 2217 |

**Declaration by Participant**

- I have read the Participant Information Sheet or someone has read it to me in a language that I understand.
- I understand the purposes, procedures and risks of the research described in the project.
- I have had an opportunity to ask questions and I am satisfied with the answers I have received.
- I freely agree to participate in this research project as described and understand that I am free to withdraw at any time during the study without affecting my future health care.
- I understand that I will be given a signed copy of this document to keep.

Name of Participant (please print) ______________________________

Signature ______________________ Date ______________________

**Declaration by Study Doctor/Senior Researcher†**

I have given a verbal explanation of the research project, its procedures and risks and I believe that the participant has understood that explanation.

Name of Surgeon/Senior Researcher†
(please print) ______________________________

Signature ______________________ Date ______________________

† A senior member of the research team must provide the explanation of, and information concerning, the research project.

Note: All parties signing the consent section must date their own signature.
